# The α_2A_-adrenergic receptor (*ADRA2A*) modulates susceptibility to Raynaud’s syndrome

**DOI:** 10.1101/2023.10.04.23296526

**Authors:** Anniina Tervi, Markus Ramste, Erik Abner, Paul Cheng, Jacqueline M. Lane, Matthew Maher, Vilma Lammi, Satu Strausz, Trieu Nguyen, Mauro Lago Docampo, Wenduo Gu, FinnGen, Estonian biobank research team, Tõnu Esko, Richa Saxena, Aarno Palotie, Samuli Ripatti, Nasa Sinnott-Armstrong, Mark Daly, Marlene Rabinovitch, Caroline A. Heckman, Thomas Quertermous, Samuel E. Jones, Hanna M. Ollila

## Abstract

Raynaud’s syndrome is a common dysautonomia where exposure to cold increases the vascular tone of distal arteries causing vasoconstriction and hypoxia, particularly in the extremities. Current treatment options are limited and unspecific. Biological mechanisms leading to the phenotype remain uncharacterized. Using genetic and electronic health record data from the UK Biobank, the Mass-General Brigham Biobank, the Estonian Biobank, and the FinnGen study, we identified 11,358 individuals with a diagnosis of Raynaud’s syndrome and 1,106,871 population controls. We found eight loci including endothelial nitric oxide synthase (*NOS3*), HLA, and a notable association at the α_2A_-adrenergic receptor (*ADRA2A)* locus (rs7090046, P = 3.93×10^-47^), implicating adrenergic signaling as a major risk factor with Raynaud’s syndrome. We further investigate the role of the variants and *ADRA2A* expression in functional and physiological models. *In silico* follow-up analysis revealed an expression quantitative trait locus (eQTL) that co-localized and increased *ADRA2A* gene expression in a tissue-specific manner in the distal arteries. Staining with RNA scope further clarified the specificity of *ADRA2A* expression in small vessels. We show by CRISPR gene editing that the SNP region modifies *ADRA2A* gene expression in pulmonary artery smooth muscle cells. Finally, we performed a functional contraction assay on smooth muscle cells in cold conditions and showed lower contraction in *ADRA2A*-deficient and higher contraction in *ADRA2A*-overexpressing smooth muscle cells. Our results indicate that Raynaud’s syndrome is related to vascular function mediated by adrenergic signaling through *ADRA2A*. Our study highlights the power of genome-wide association testing as a discovery tool for poorly understood clinical endpoints and further clarifies the role of adrenergic signaling in Raynaud’s syndrome by fine-mapping, using *in vitro* genomic manipulations and functional validation in distal smooth muscle cell populations located in arterioles

## Main

The autonomic nervous system controls physiological functions in the body that are not under direct voluntary control and are not typically consciously directed. The targets of the autonomic nervous system include body temperature, heart rate, respiration, bowel movements and digestion, sexual arousal, endocrine function, blood pressure regulation and vascular tone. However, when the autonomic nervous system malfunctions, it can lead to symptoms and diseases of dysautonomia, affecting many different functions of the autonomic nervous system including vascular tone and blood pressure as seen with Raynaud’s syndrome (RS)^1^.

RS has a clear and specific disease manifestation, where the exposure to cold increases the vascular tone of distal arteries causing vasoconstriction, leading to cyanosis and hypoxia particularly in fingers and toes^2^. RS can be seen as an example of a disease with clear component of dysautonomia. Furthermore, RS is a common phenomenon, with an estimated prevalence of 3 to 5% in the global population^3^. RS rarely causes clinically debilitating symptoms, but RS is diagnosed with a single code in the international classification of diseases (ICD-10 I73.0^4^) making it possible to use electronic health records to find individuals with clinically significant RS and consequently understand RS disease manifestation, disease correlations and the underlying biological mechanisms.

Moreover, the comorbidities of RS include symptoms of pulmonary hypertension in a subset of patients, especially in patients with systemic sclerosis^5-8^. Consequently, RS can manifest as a comorbidity of diseases with substantial clinical significance, such as systemic sclerosis, lupus erythematosus, myalgic encephalomyelitis/chronic fatigue syndrome, and most recently Long COVID^9-11^. Elucidating the disease mechanisms behind primary RS would potentially provide insight into diseases with dysautonomia^12^.

Finally, RS has a relatively high hereditary component with estimated twin heritability between 54-65%^13,14^. This study is the first to examine genetic associations of RS across multiple cohorts and with functional validation in cellular models.

## Results

### α_2A_-adrenergic receptor associates with Raynaud’s syndrome

Using genetic and electronic health record data from FinnGen data freeze 10 (R10), the UK Biobank (UKB), the Estonian Biobank (EstBB), and the Mass General Brigham Biobank (MGB), we identified a total of 11,358 individuals with diagnosis of RS and 1,106,871 controls. Demographic characteristics of in the study cohorts showed that the majority of RS patients were females (73.2 %), in agreement with earlier reports^15,3^, and mean disease diagnosis age was at 49.6 years (Suppl. Table 1).

Genome-wide association analysis of RS identified eight loci, which included a notable association at the α_2A_-adrenergic receptor (*ADRA2A*) locus, which was seen independently at genome-wide significant level (P ≤ 5×10^-8^) in all four cohorts and was further supported in the meta-analysis (rs7090046, P meta-analysis = 3.93×10^-47^, beta [SE] = 0.22 [0.02], Figure 1, Table 1, Suppl. Figs. 1&2, Suppl. Tables 2,3,4).

**Figure 1.**
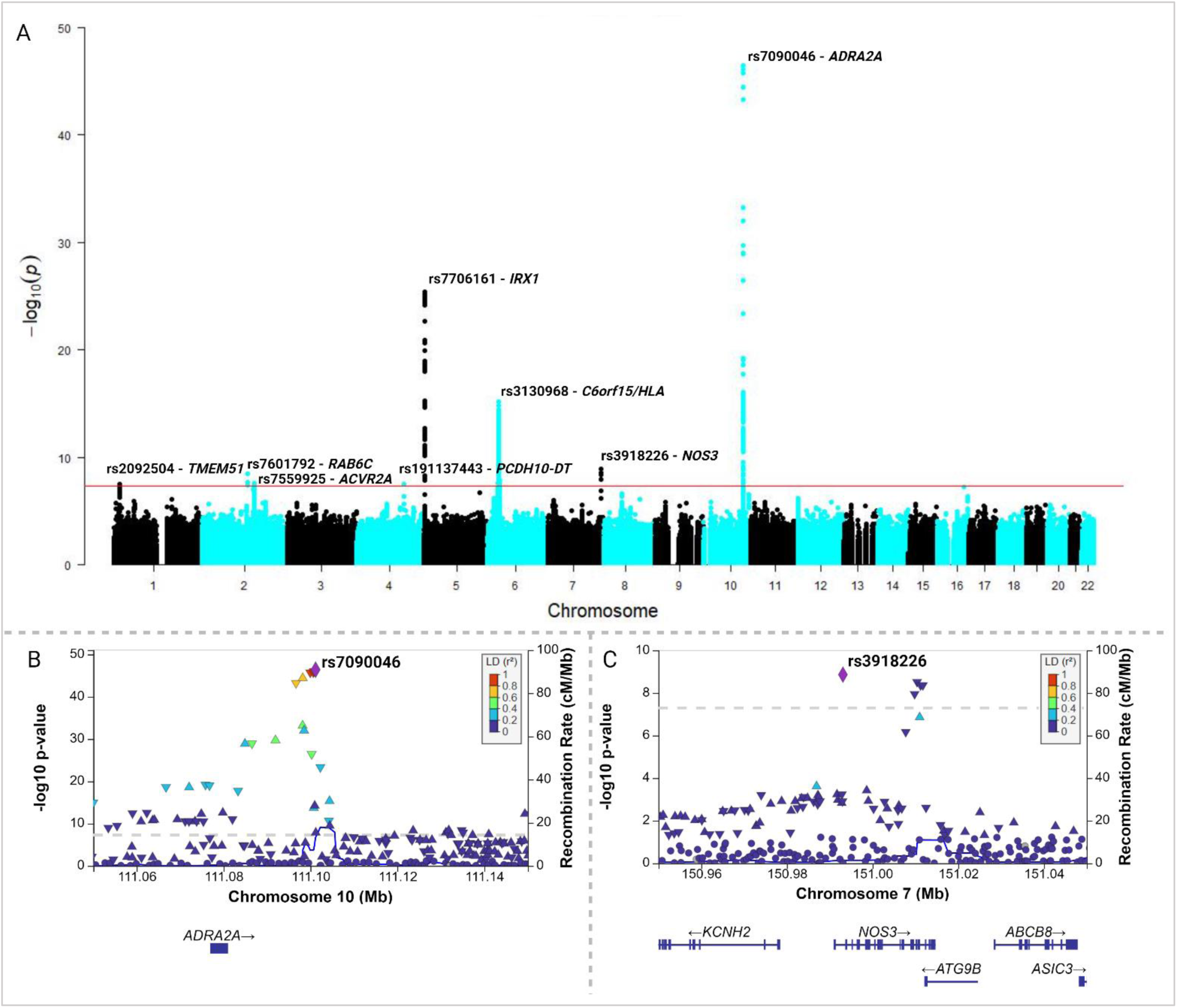
Meta-analysis. **A.** A Manhattan plot of RS meta-analysis combining UKB, FinnGen R10, MGB and EstBB. **B.** Locus zoom plots for regional association at the *ADRA2A* locus, lead variant rs7090046 and **C.** regional association at the *NOS3* locus, lead variant rs3918226. (Created with Biorender.com)

**Table 1.**
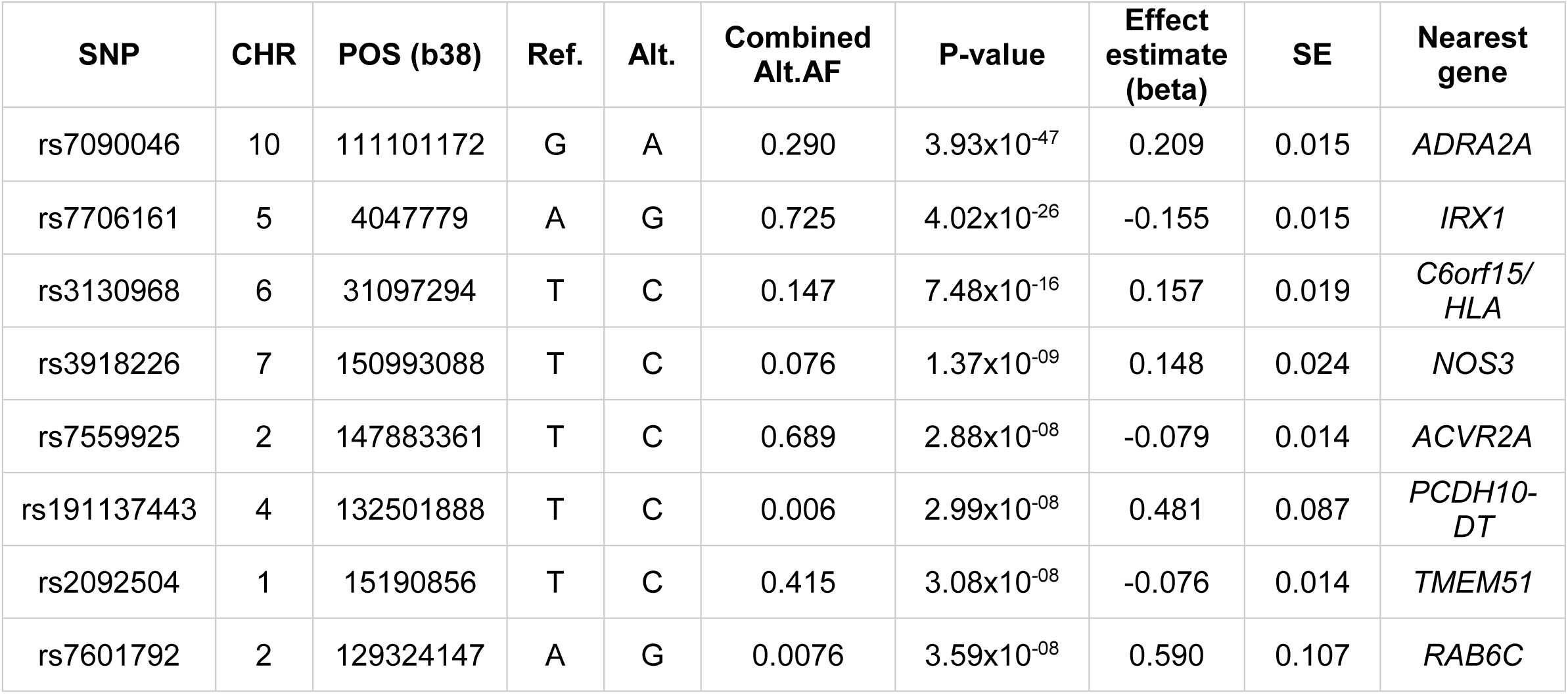
RS meta-analysis lead variants combining the UKB, FinnGen R10, MGB and EstBB data.

| SNP | CHR | POS (b38) | Ref. | Alt. | Combined Alt.AF | P-value | Effect estimate (beta) | SE | Nearest gene |
| --- | --- | --- | --- | --- | --- | --- | --- | --- | --- |
| rs7090046 | 10 | 111101172 | G | A | 0.290 | $3.93 \times 10^{-47}$ | 0.209 | 0.015 | *ADRA2A* |
| rs7706161 | 5 | 4047779 | A | G | 0.725 | $4.02 \times 10^{-26}$ | -0.155 | 0.015 | *IRX1* |
| rs3130968 | 6 | 31097294 | T | C | 0.147 | $7.48 \times 10^{-16}$ | 0.157 | 0.019 | *C6orf15/HLA* |
| rs3918226 | 7 | 150993088 | T | C | 0.076 | $1.37 \times 10^{-9}$ | 0.148 | 0.024 | *NOS3* |
| rs7559925 | 2 | 147883361 | T | C | 0.689 | $2.88 \times 10^{-8}$ | -0.079 | 0.014 | *ACVR2A* |
| rs191137443 | 4 | 132501888 | T | C | 0.006 | $2.99 \times 10^{-8}$ | 0.481 | 0.087 | *PCDH10-DT* |
| rs2092504 | 1 | 15190856 | T | C | 0.415 | $3.08 \times 10^{-8}$ | -0.076 | 0.014 | *TMEM51* |
| rs7601792 | 2 | 129324147 | A | G | 0.0076 | $3.59 \times 10^{-8}$ | 0.590 | 0.107 | *RAB6C* |

In our meta-analysis, additional seven loci were genome-wide significant (P ≤ 5×10^-8^, Table 1). The variant rs7706161 is an intergenic variant closest to and downstream from *IRX1* (iroquois homeobox 1) gene. *IRX1* encodes for a homeobox gene involved in finger development in model organisms^16^. Furthermore, rs3130968 at chromosome 6 is located in the HLA region downstream from *HCG22* (HLA complex group 22) and upstream from the *HLA-C* and *HLA-B* genes. The same variant has also been associated with peripheral vascular disease^17^. Moreover, the lead genome-wide significant variant in chromosome 7, rs3918226, is an intron variant for *NOS3* (nitric oxide synthase 3/endothelial nitric oxide) 5’ regulatory region. As endothelial nitric oxide signaling (eNOS) is a canonical mechanism in vasoconstriction and dilation^18,19,20^, our findings indicate a role for eNOS as part of vasoconstriction also in RS.

Variant rs7601684 in chromosome 2 is located upstream from *RAB6C,* a member of the RAS oncogene family. The other chromosome 2 variant, rs7559925, is an intron variant for the *ACVR2A* gene (activin A receptor type 2A) that encodes for a receptor related to activins, which are part of the TGF-beta (transforming growth factor-beta) family of proteins. Additionally, variant rs2092504 in chromosome 1, an intronic variant in transmembrane protein 51 (*TMEM51*), and variant rs191137443 in chromosome 4 upstream from protocadherin 10 gene (*PCDH10*) were genome-wide significant in our meta-analysis. The association with rs7559925 was shown with immunological traits such as lymphocyte counts^21^ whereas rs3918226 has been shown to be associated with high blood pressure^22^.

While many of these variants discovered in this GWAS are related to vascular tone, blood pressure or immune function, the *ADRA2A* locus stands out as a significant association in all tested cohorts and biobanks, highlighting its importance in RS across multiple population cohorts. *ADRA2A* encodes for the α_2A_-adrenergic receptor, is targeted by α-blockers, and has a downstream effect on lowering vascular tone and consequently blood pressure^23-28^.

### *ADRA2A* RS risk variants increase *ADRA2A* expression

To understand the functional consequences of the *ADRA2A* variants, we examined their association with gene expression across human tissues and tissue specificity. Using data from GTEx (https://gtexportal.org/home/), we observed that the lead variant (rs7090046) affected the expression of *ADRA2A* in a tissue-specific manner in the tibial arteries (rs7090046, eQTL P = 1.3×10^-13^, Figure 2) in contrast to coronary arteries (P = 0.16) or aorta (P = 0.65). In addition, the lead variant for *ADRA2A* expression in GTEx in tibial arteries was rs1343449 (r^2^ in Europeans with rs7090046 = 0.98), which was also the lead variant in the MGB and among the five top variants in each cohort and the credible set (Figure 2, Suppl. Table 2). A formal co-localization analysis suggested a shared signal between RS and *ADRA2A* expression in tibial arteries specifically (posterior probability = 0.99, Figure 2, Suppl. Fig. 3, Suppl. Table 7). The risk allele associated with higher RS risk associated with higher *ADRA2A* expression in agreement with earlier role of adrenergic receptors regulating vascular wall contraction^29^. Finally, we performed stratified LD score regression using ENCODE^30^ to elucidate overall relevance of different tissues across the body in RS. We discovered the most significant associations with smooth muscle cell (SMC) types, further suggesting that possible pathology is mediated by SMCs (Suppl. Table 5).

**Figure 2.**
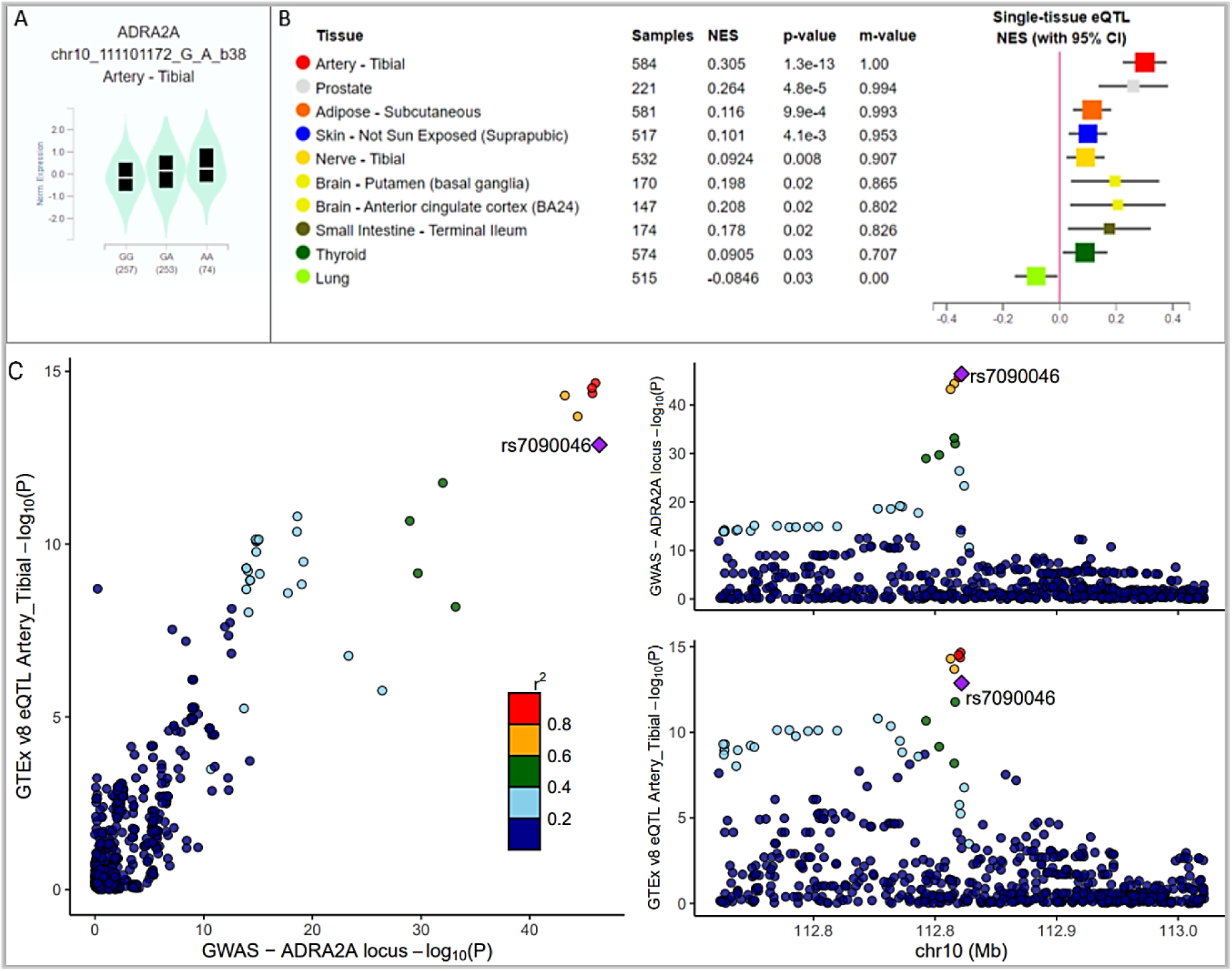
*ADRA2A* expression. **A.** Genotype-tissue expression for SNP rs7090046 in tibial arteries) and **B.** across tissues from GTEx (NES = normalized expression values, m-value = posterior probability, P ≤ 0.05. **C.** the RS association colocalizes with eQTL signal in tibial arteries. (created with Biorender.com)

### *ADRA2A* is specifically expressed in microvasculature

Human arteries contain primarily fibroblasts, SMCs, pericytes (defined as cells found around microvasculature), and endothelial cells. As *ADRA2A* eQTL suggests that the variant affects expression in blood vessels in particular, we wanted to examine which cell types express *ADRA2A* in the vascular wall. First, single cell RNA sequencing (scRNAseq) data from human vascular tissue demonstrated that only a small cluster of cells expresses *ADRA2A* (Figure 3B). This cluster expressing *ADRA2A* is distinct from the medial smooth muscle cells (SMCs) that are the majority of the *MYH11*+, a classical marker for medial smooth muscle cells that are found in the vessel wall (Figure 3B & 3D). Moreover, the cluster expressing *ADRA2A* contains cells that express mature SMC genes, and in addition co-express *NOTCH3* (Figure 3B & 3C)^31,32^. These findings suggest that only microvascular cells express *ADRA2A* and are likely a causal cell population for RS.

**Figure 3.**
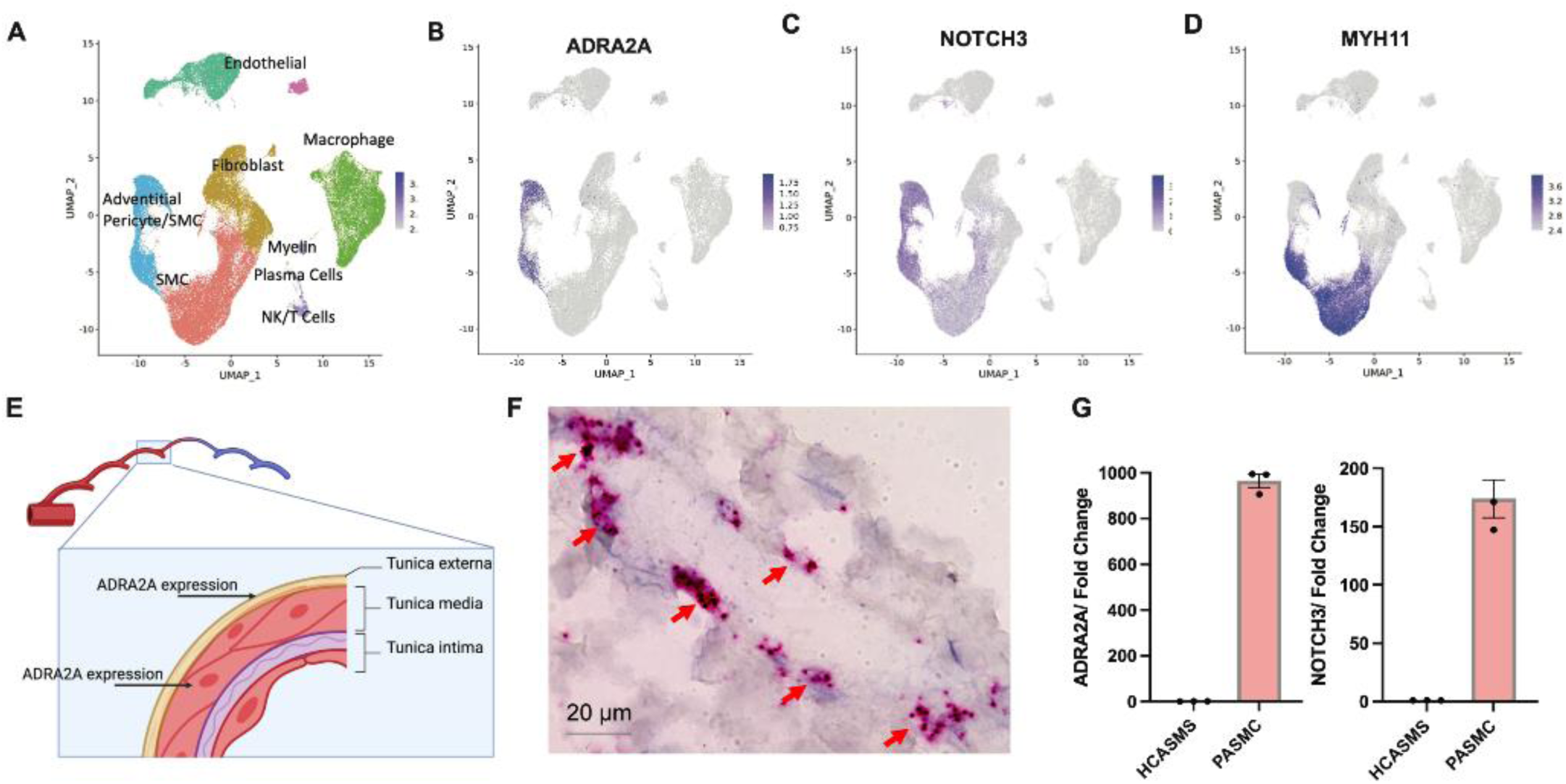
*ADRA2A* expression is restricted to microvascular SMC. UMAP plot of human artery scRNAseq from human vascular atlas indicating **A.** cell type identities of all clusters, B. *ADRA2A* expression, **C.** Co-expression of *NOTCH3* in the same cluster **D.** and *MYH11* expression. **E.** Schematic of vascular wall and *ADRA2A* expression. **F.** RNA Scope against *ADRA2A* in distal human coronary arteries **G.** RT-PCR quantification of *ADRA2A* and *NOTCH3* expression HCASMs and PASMCs.

To further understand the role of the *ADRA2A* expressing cell population, we assessed the expression of *ADRA2A* in 14 different primary vascular smooth muscle cells. *ADRA2A* was found in only one line that was likely obtained from the pulmonary microvasculature but *ADRA2A* expression was otherwise not detected in any other vascular SMC lines derived from larger arteries including medium to large arteries (Figure 3F & 3G). This particular cell line also expresses other markers of the microvascular SMC population identified by scRNAseq, such as *NOTCH3* (Figure 3G). Thus, this microvascular SMC line was chosen for subsequent functional studies.

### *ADRA2A* expression affects SMC contraction in temperature dependent fashion

Next, we examined how *ADRA2A* expression alters SMC contractility in conditions mimicking RS and environmental cold stress. Earlier studies in temperature-dependent vascular contraction have supported the role of the adrenergic system but have focused nearly solely on the role of *ADRA2C* and its temperature-dependent activation^26,33,34^. These earlier studies and our findings raise an interesting question: Does *ADRA2A* directly affect vascular contraction? To test this, we used a collagen-based SMC contraction assay in *ADRA2A* over-expressing or silenced cells. We discovered that in cold conditions (+28°C), silenced siRNA-*ADRA2A* treated pulmonary artery SMCs (PASMCs) contracted significantly less (Figure 4A). However, *ADRA2C* silencing did not attenuate cold induced contraction in a similar manner to *ADRA2A* (Figure 4A). In warm conditions, both *ADRA2A* and *ADRA2C* silenced SMCs contracted similarly (Figure 4B). Consequently, upon lentiviral overexpression of *ADRA2A*, we observed that the PASMCs contracted significantly more (Figure 4C, D) overall suggesting that *ADRA2A* was affecting contraction in a dose dependent fashion and was responsible for cold induced contraction.

**Figure 4.**
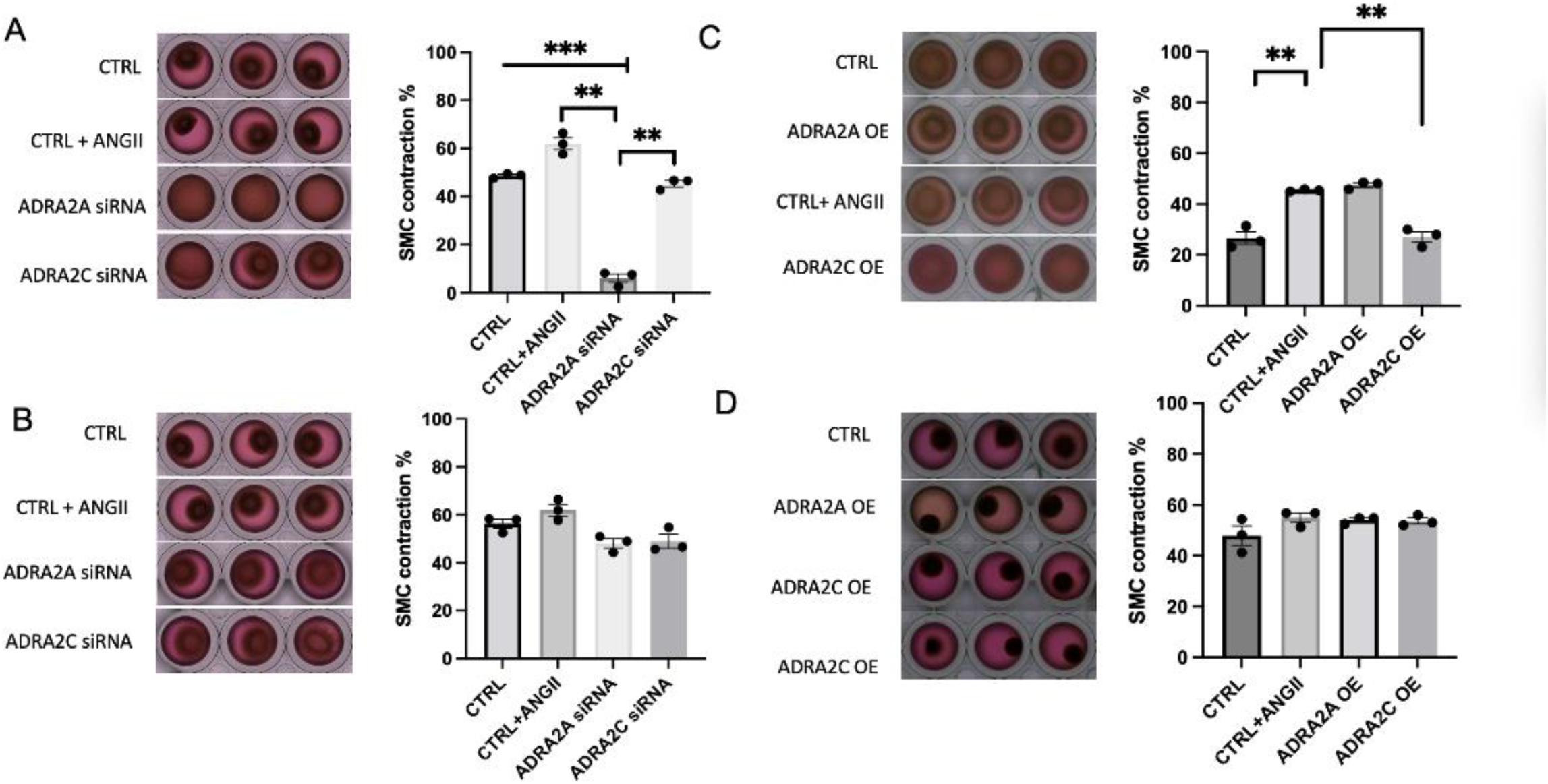
ADRA2A expression affects SMC contraction upon cold stimulus (+28°C). **A.** *ADRA2A* and *ADRA2C* silencing in cold exposure **B.** *ADRA2A* and *ADRA2C* silencing and in ambient conditions **C.** *ADRA2A* and *ADRA2C* overexpression in cold exposure **D.** *ADRA2A* and *ADRA2C* overexpression in ambient conditions. Mean +/- SEM, ***P<0.0005, **P<0.005 & *P<0.05.

### CRISPR interference targeting rs7090046 supports *ADRA2A’s role* as a causal gene for RS

We then used this cell line to study *ADRA2A* gene causality in RS. To elucidate mechanistic importance of the *ADRA2A* rs7090046 locus, we designed five CRISPR guides targeting the lead SNP variant rs7090046 and used the CRISPRi-Cas9 machinery to interfere signaling from this variant region. We observed a significant decrease in *ADRA2A* gene expression 5 days after the lentiviral treatment on the PASMCs suggesting that the region is needed for controlling *ADRA2A* expression (Figure 5).

**Figure 5.**
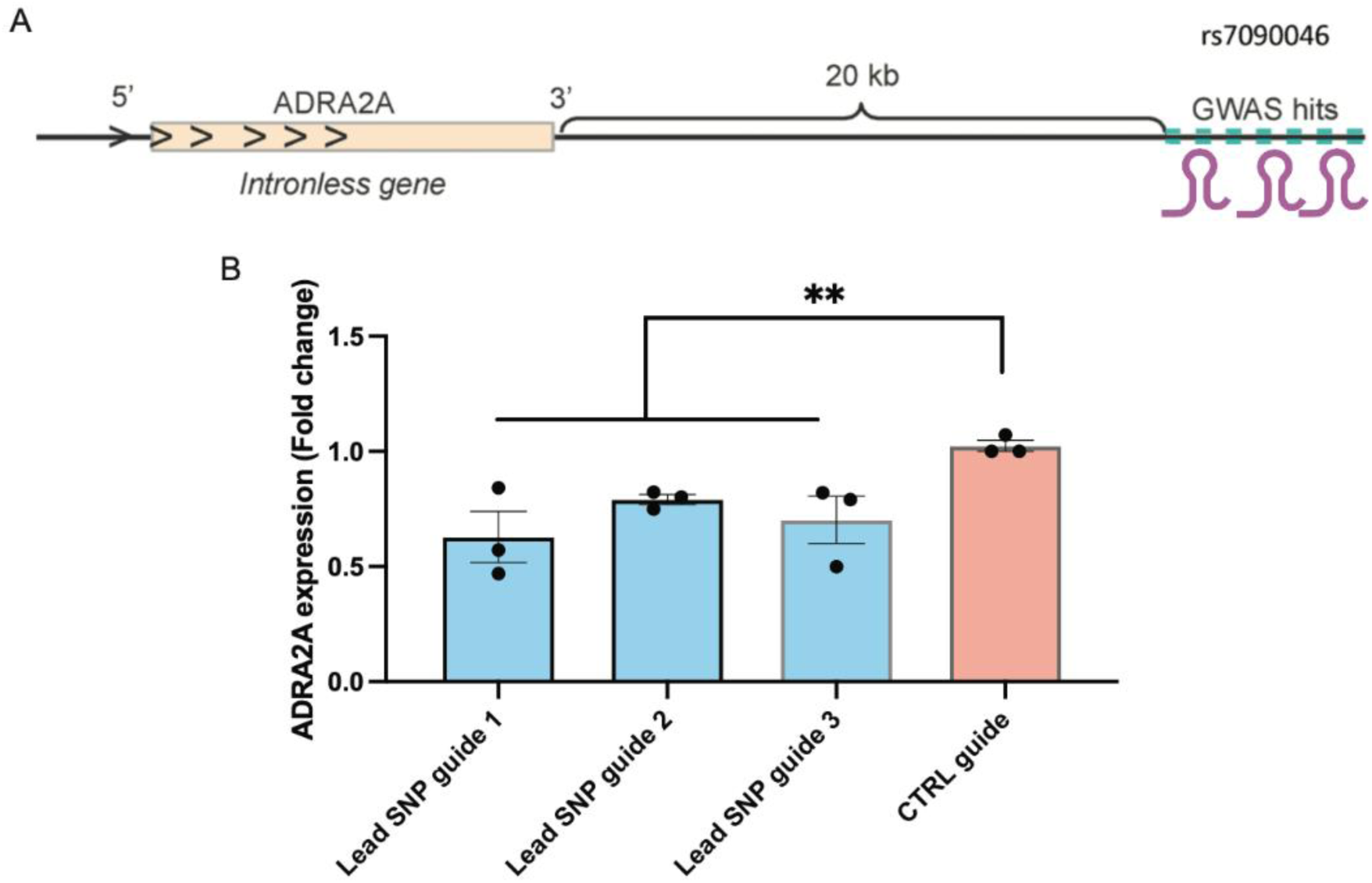
CRISPR interference against rs7090046 supports causal role of *ADRA2A*. **A.** Schematic of the CRISPR interference experiment. **B.** RT-PCR quantification of *ADRA2A* expression in rs7090046 guide targeted cells vs CTRL. Mean +/- SEM, ***P<0.0005, **P<0.005 & *P<0.05.

## Discussion

We performed a meta-analysis of RS across four cohorts and identified genome-wide significant associations with RS at *ADRA2A, HLA, NOS3, RAB6C, ACVR2A, PCDH10, TMEM51* and *IRX1* loci. The most prominent genetic association with RS was discovered in the *ADRA2A* locus. Furthermore, this association was significant independently in all cohorts, highlighting the significance of *ADRA2A* in RS. *In silico* follow-up analysis of *ADRA2A* RNA expression across tissues showed the highest expression in tibial arteries, and single cell expression analysis further supported the role of SMCs as the key cell type for *ADRA2A* expression. Finally, functional contraction assay in SMCs in cold conditions showed lower contraction in *ADRA2A*-deficient and higher contraction in *ADRA2A* overexpressing SMCs. Overall, our findings indicate *ADRA2A* in RS and as a regulator of vascular contraction in SMCs in temperature dependent fashion.

The strongest association in this study is a robust signal from *ADRA2A* in all four cohorts and with the same lead signals across cohorts. Furthermore, the lead variants are located in a regulatory region that affected *ADRA2A* expression in distal arteries in particular. In addition, we identified a specific subpopulation of SMCs that specifically expresses the α_2A_-adrenergic receptor. While the adrenergic system has been suggested as a potential pathological mechanism underlying RS^23,24,26,27,33-41^, these earlier studies have focused almost solely on the α_2C_-adrenergic receptor. Interestingly, a recent biobank study has identified some of the same RS loci^42^. In this study, we assessed contraction SMCs after *ADRA2A* or *ADRA2C* siRNA knockdown in human SMCs. We saw a robust contraction upon cold with *ADRA2A* knockdown whereas *ADRA2C* silencing did not attenuate cold induced contraction in a similar manner to *ADRA2A*. Overall, these findings suggest an independent role of *ADRA2A* in vascular contraction and in temperature-dependent control of vascular tone.

In cold or stress conditions, norepinephrine and epinephrine are released and bind to adrenergic receptors throughout the body, which exerts various effects: dilating pupils and bronchioles, increase in heart rate, and in blood vessels constriction. The blood vessels constriction is mediated by the SMC that express adrenergic receptors on their surface. In healthy patients without RS, there are mechanisms to prevent unwanted and excessive vessel contraction. First, the cell constriction (upon cold or stress) can be limited by the increased release of ligand by translocation of the α_2C_-adrenergic receptor from the cytosol to the cell surface. Second, α_2A_-adrenergic receptors are also expressed on the presynaptic membrane and function there as a negative feedback loop for catecholamine release^43^.

Based on our results we propose a new model explaining the pathomechanism of RS that underlines the power of human genetics driven studies for understanding disease mechanisms. Our SMC contraction assays show that cold induced SMC contractility was modified by *ADRA2A* expression. Furthermore, we combined functional studies with the genomics-driven discovery that genetic variation at the *ADRA2A* locus leads to increased expression of α_2A_-adrenergic receptor in a population of SMCs. Such an increase in *ADRA2A* expression may sensitize these cells to adrenergic response and lead to increased signaling through the adrenergic pathway (Figure 6).

**Figure 6.**
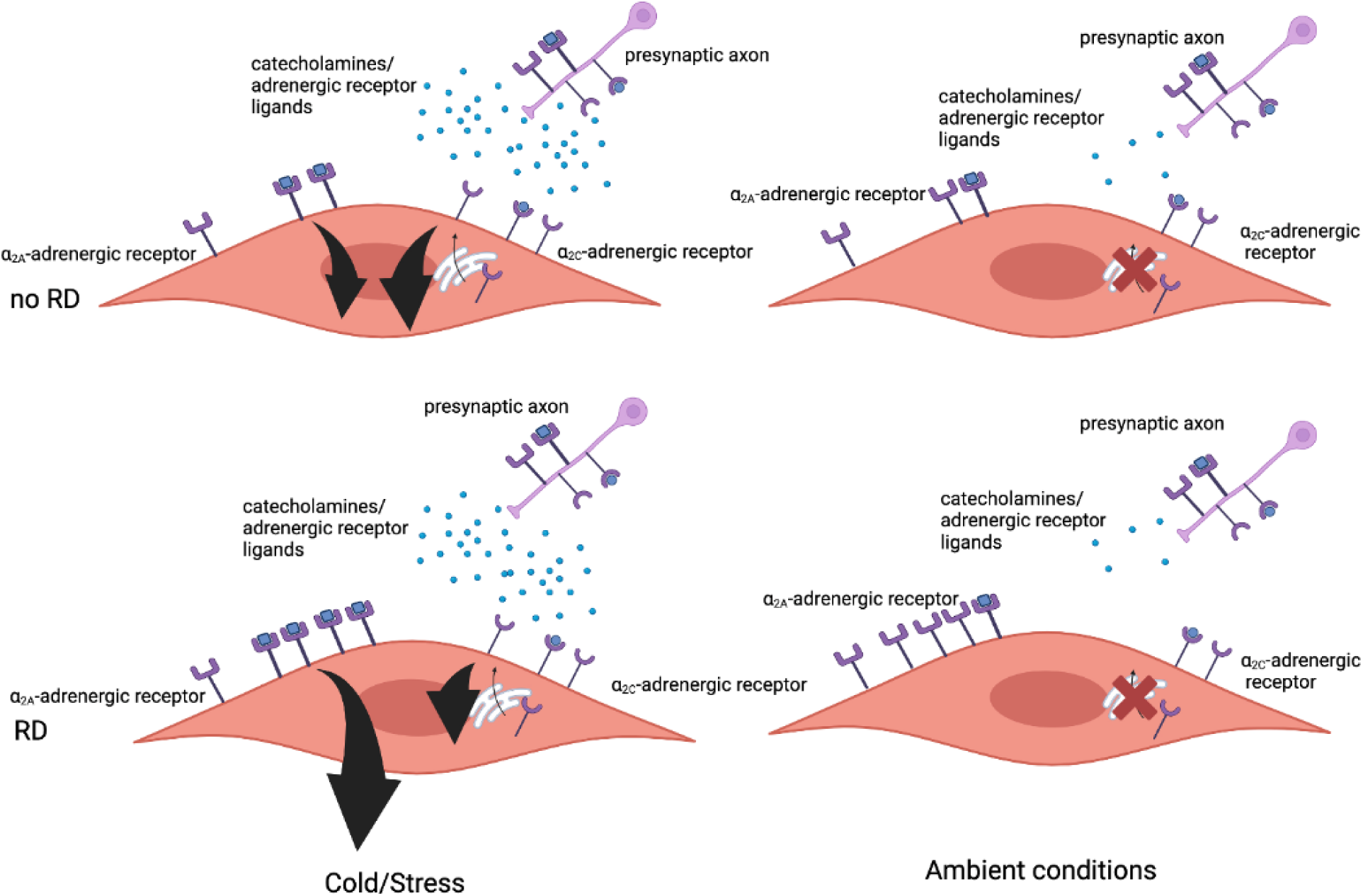
Schematic of the proposed RS –associated pathomechanism. We propose a possible mechanism based on our observations from genetic and contraction assays. In this hypothesized model, the pathomechanism of RS is dependent on *ADRA2A* expression. In normal physiological conditions, it is well established that cold or stress both induce secretion of the ligands that bind the adrenergic receptors. Consequently, these ligands such as epinephrine and norepinephrine activate the adrenergic system. In RS patients, the expression of *ADRA2A* gene encoding α2A-adrenergic receptor is higher and similarly the amount of α2A-adrenergic receptor available for ligand binding on microvascular SMCs. Increase in SMC *ADRA2A* expression sensitizes the postsynaptic system that aggravates adrenergic effects of SMC contraction. In conditions such as cold and stress where more ligand is released the contraction is further accentuated. Black arrows representing adrenergic downstream signaling strength.

In addition to the signal from the adrenergic system, our meta-analysis indicated other signals and in particular endothelial nitric oxide synthase (eNOS) that clarify the pathological mechanisms with RS. Nitric oxide itself is a well-known mediator of vasodilation in arteries^44^. Therefore, it has been suggested as one of the underlying pathological mechanisms in RS although the role of it is still unclear^2,45,46^. These genetic associations may also elucidate the more specific disease mechanisms or even provide insight into the heterogeneity of the symptomatology in RS. For example, the signal from chromosome 6, HLA class I region, points towards the possible immune, infectious or autoimmune mechanisms in RS. It is still unclear, however, if this signal is a signal from autoimmune diseases that have secondary RS and not primary Raynaud’s disease per se^47,48^, which was also indicated by Hartmann *et al.* (2023) in the UKB^42^. As for the other signals detected by our meta-analysis, *IRX1* gene has been implied in the embryonic development and in the development of fingers and digits which are affected in RS^49,50^ as well as a tumor suppression gene^51-54^. Variants in *ACVR2A, RAB6C, PCDH10* and *TMEM51* have been implied in immunological traits, cell cycle, neurological disease and contractile function in cardio myocytes, respectively^21,55-57^.

Our study should be interpreted within the following limitations. Only a subset of individuals with RS have symptoms that are recognized or need treatment and we are likely missing the more benign spectrum of symptoms in the current population. Our findings are limited to individuals with European ancestry. Finally, data from subcutaneous microvascular cells or microvascular cells from RS patients were not available for this study and any genetic or expression effects might be accentuated or changed after disease onset in RS patients.

To summarize, we report here a robust association with *ADRA2A* with functional follow-up of this locus. In addition, we discovered seven loci that can be followed up in future functional studies. Our findings not only point towards the vascular pathophysiology underlying RS, but also specifically to the dysfunction of the autonomic nervous system and its dialogue with the vascular structure. Our results also suggest that the pathophysiological RS phenotype can be mediated through multiple possibly additive mechanisms involving adrenergic signaling, immune mechanisms and nitric oxide action and are in agreement with earlier studies. We highlight *ADRA2A* in the adrenergic system as a likely causal gene in a subset of microvascular SMCs, where we suggest it sensitizes these cells for catecholamines. Due to its localized expression in this subset of cells and the dose-dependent nature of the pathological phenotype as demonstrated by the genetic data, *ADRA2A* may be a promising target for further pharmaceutical studies.

## Materials and Methods

### Genetic analyses

#### Cohorts

FinnGen is a public-private partnership registry-based study of Finnish residents combining genetic and electronic health record data form different registers, for example, primary care and hospital in- and out-patient visits. The release 10 (R10) contains data on up to 412,181 participants, primarily of Finnish ancestry from newborns to the age of 104 at baseline recruitment. The aim of the study is to collect the data of 500,000 Finns representing 10% of the population of Finland (for more information see https://www.finngen.fi/en).

The UK Biobank (UKB) is a population-based study containing over 500,000 individuals of mainly European ancestry^58^. The participants were recruited to the study between 2006 to 2010, were aged between 37 to 73 years of age and were residents of the United Kingdom. The study is a combination of, for example, different lifestyle measures, genotypes, electronic health record data, blood count data and questionnaire data, and the data is updated frequently to capture the health trajectories of participated individuals.

The Estonian Biobank is a population-based biobank with 212,955 participants in the current data freeze (2023v1). All biobank participants have signed a broad informed consent form and information on ICD-10 codes is obtained via regular linking with the national Health Insurance Fund and other relevant databases, with majority of the electronic health records having been collected since 2004^59^.

The Mass-General Brigham (MGB) Biobank (formerly Partners HealthCare Biobank) is a hospital-based cohort study from the MGB healthcare network in Boston (MA, USA) with electronic health record (EHR), genetic, and lifestyle data^60-62^. The MGB Biobank includes data obtained from patients in several community-based primary care facilities and specialty tertiary care centers in Boston, MA^60,63^.

#### Ethics statement

Patients and control subjects in FinnGen provided informed consent for biobank research, based on the Finnish Biobank Act. Alternatively, separate research cohorts, collected prior the Finnish Biobank Act came into effect (in September 2013) and start of FinnGen (August 2017), were collected based on study-specific consents and later transferred to the Finnish biobanks after approval by Fimea (Finnish Medicines Agency), the National Supervisory Authority for Welfare and Health. Recruitment protocols followed the biobank protocols approved by Fimea. The Coordinating Ethics Committee of the Hospital District of Helsinki and Uusimaa (HUS) statement number for the FinnGen study is Nr HUS/990/2017.

The FinnGen study is approved by Finnish Institute for Health and Welfare (permit numbers: THL/2031/6.02.00/2017, THL/1101/5.05.00/2017, THL/341/6.02.00/2018, THL/2222/6.02.00/2018,THL/283/6.02.00/2019, THL/1721/5.05.00/2019 and THL/1524/5.05.00/2020), Digital and population data service agency (permit numbers: VRK43431/2017-3, VRK/6909/2018-3, VRK/4415/2019-3), the Social Insurance Institution (permit numbers: KELA 58/522/2017, KELA 131/522/2018,KELA 70/522/2019, KELA 98/522/2019, KELA 134/522/2019, KELA 138/522/2019, KELA 2/522/2020,KELA 16/522/2020), Findata permit numbers THL/2364/14.02/2020, THL/4055/14.06.00/2020, THL/3433/14.06.00/2020, THL/4432/14.06/2020, THL/5189/14.06/2020, THL/5894/14.06.00/2020, THL/6619/14.06.00/2020, THL/209/14.06.00/2021, THL/688/14.06.00/2021, THL/1284/14.06.00/2021,THL/1965/14.06.00/2021, THL/5546/14.02.00/2020 and Statistics Finland (permit numbers: TK-53-1041-17 and TK/143/07.03.00/2020 (earlier TK-53-90-20)).

The Biobank Access Decisions for FinnGen samples and data utilized in FinnGen Data Freeze7 include: THL Biobank BB2017_55, BB2017_111, BB2018_19, BB_2018_34, BB_2018_67, BB2018_71, BB2019_7, BB2019_8, BB2019_26, BB2020_1, Finnish Red Cross Blood Service Biobank 7.12.2017, Helsinki Biobank HUS/359/2017, Auria Biobank AB17-5154 and amendment #1 (August 17 2020), Biobank Borealis of Northern Finland_2017_1013, Biobank of Eastern Finland 1186/2018 and amendment 22 § /2020, Finnish Clinical Biobank Tampere MH0004 and amendments (21.02.2020 06.10.2020), Central Finland Biobank 1-2017, and Terveystalo Biobank STB 2018001.2.

The activities of the EstBB are regulated by the Human Genes Research Act, which was adopted in 2000 specifically for the operations of the EstBB. Individual level data analysis in the EstBB was carried out under ethical approval 1.1-12/624 from the Estonian Committee on Bioethics and Human Research (Estonian Ministry of Social Affairs), using data according to release application 6-7/GI/16279 from the Estonian Biobank.

The North West Multi-centre Research Ethics Committee (MREC) has granted the Research Tissue Bank (RTB) approval for the UKB that covers the collection and distribution of data and samples (http://www.ukbiobank.ac.uk/ethics/). Our work was performed under the UKB application number 22627 (Principal Investigator Dr Matti Pirinen, FIMM). All participants included in the conducted analyses have given a written consent to participate.

The MGB has obtained a Certificate of Confidentiality. In addition, The MGB works in close collaboration with the Partners Human Research Committee (PHRC) (the Institutional Review Board). This collaboration has ensured that the Biobank’s actions and procedures meet the ethical standards for research with human subjects. Biobank patients are recruited from inpatient stays, emergency department settings, outpatient visits, and electronically through a secure online portal for patients. Recruitment and consent materials are fully translated in Spanish to promote patient inclusion. The systematic enrollment of patients across the MGB network and the active inclusion of patients from diverse backgrounds contribute to a Biobank reflective of the overall demographic of the population receiving care within the MGB network. Recruitment for the Biobank launched in 2009 and is ongoing through both in-person recruitment at participating clinics and electronically through the patient portal. The recruitment strategy has been described previously^60^. All recruited patients provided written consent upon enrollment, and are offered an option to refuse consent.

#### Genotyping and quality control

Genotyping in the FinnGen cohort was performed by using Illumina (Illumina Inc., San Diego, CA, USA) and Affymetrix arrays (Thermo Fisher Scientific, Santa Clara, CA, USA) and lifted over to build version 38 (GRCh38/hg38)^64^. Individuals with high genotype absence (> 5%), inexplicit sex or excess heterozygosity (+-4 standard deviations) were excluded from the data^64^. Additionally, variants that had high absence (> 2%), low minor allele count (< 3) or low Hardy-Weinberg Equilibrium (HWE) (P < 1×10^-06^) were removed. More detailed explanations of the genotyping, quality control and the genotype imputation are provided elsewhere^64^. All individuals in the cohort were Finns and matched against the SiSu v4 reference panel (http://www.sisuproject.fi/).

All the EstBB participants have been genotyped at the Core Genotyping Lab of the Institute of Genomics, University of Tartu, using Illumina Global Screening Array v3.0_EST. Samples were genotyped and PLINK format files were created using Illumina GenomeStudio v2.0.4. Individuals were excluded from the analysis if their call-rate was < 95%, if they were outliers of the absolute value of heterozygosity (> 3SD from the mean) or if sex defined based on heterozygosity of X chromosome did not match sex in phenotype data. Before imputation, variants were filtered by call-rate < 95%, HWE P-value < 1×10^-04^ (autosomal variants only), and minor allele frequency < 1%. Genotyped variant positions were in build 37 and were lifted over to build 38 using Picard. Phasing was performed using the Beagle v5.4 software^65^. Imputation was performed with Beagle v5.4 software (beagle.22Jul22.46e.jar) and default settings. Dataset was split into batches 5,000. A population specific reference panel consisting of 2,695 WGS samples was utilized for imputation and standard Beagle hg38 recombination maps were used. Based on principal component analysis, samples who were not of European ancestry were removed. Duplicate and monozygous twin detection was performed with KING 2.2.7^66^, and one sample was removed out of the pair of duplicates. Analyses were restricted to individuals with European ancestry.

The UKB genotyped 488,477 participants: 49,950 on the Affymetrix (Thermo Fisher Scientific) UK BiLEVE Axiom Array and 438,427 on the highly similar Affymetrix UK Biobank Axiom Array, These arrays captured up to 825,927 SNPs and short indels, with variants prioritized for known coding variants, those previously associated with disease and ancestry-specific markers that provide a good imputation backbone. DNA was extracted from blood samples taken at baseline interviews, between 2006 and 2010, and genotyping was carried out in 106 sequential batches, giving genotype calls for 812,428 unique variants in 489,212 participants. After removing high missingness and very rare variants, as well as poor-quality samples, these genotypes were phased using SHAPEIT3 and imputed to the Haplotype Reference Consortium (HRC) and to a merged UK10K and 1000 Genomes phase 3 reference panel^67^, both in genome assembly GRCh37 using IMPUTE2. This resulted in 93,095,623 autosomal SNPs, short indels and large structural variants in 487,442 individuals and 3,963,705 markers on the X chromosome. For more details, see Bycroft *et al.* 2018^58^.

The MGB genotyped 53,297 participants on the Illumina Global Screening Array (’GSA’) and 11,864 on Illumina Multi-Ethnic Global Array ("MEG"). The GSA arrays captured approximately 652K SNPs and short indels, while the MEG arrays captured approximately 1.38M SNPs and short indels. These genotypes were filtered for high missingness (> 2%) and variants out of HWE (P < 1×10^-12^), as well as variants with an AF discordant (P < 1×10^-150^) from a synthesized AF calculated from GnomAD subpopulation frequencies and a genome wide GnomAD model fit of the entire cohort. This resulted in approximately 620K variants for GSA and 1.15M for MEG. The two sets of genotypes were then separately phased and imputed on the TOPMed imputation server (Minimac4 algorithm) using the TOPMed r2 reference panel. The resultant imputation sets were both filtered at an R2 > 0.4 and a MAF > 0.001, and then the two sets were merged/intersected resulting in approximately 19.5M GRCh38 autosomal variants. The sample set for analysis here was then restricted to just those classified as EUR (N = 54,452) according to a metric of being +/- 2 SDs of the average EUR sample’s principal components 1 to 4 in the HGDP reference panel.

#### Phenotype definition

We built the phenotype for RS using ICD10 code I73.0 in all of our cohorts (see Suppl. Table 1 for the total number of cases and controls in each cohort used). Additionally, we used the self-reported measures and primary care codes of RS in the UKB, and the ICD-9 code 4430 in the FinnGen R10 and the UKB.

For FinnGen R10 phenotype definition, we used both the hospital record data (inpatient N = 88 (4.2%) cases and outpatient N = 1,204 (57.8%) cases) and primary care data (N = 792 (38.0%)). Most of the cases (N = 2,025 (87.75%)) in the EstBB were from primary care data setting, and the rest were from hospital record data (N = 180 (12.25%)). In the case of MGB, all the cases were obtained from hospital record data (inpatient N = 251 (13.2%) cases and outpatient N = 1,656 (86.8%) cases).

From the UKB data, we obtained both self-reported and electronic health record data for disease definitions. To define the phenotypes, we used data from the self-report non-cancer illness codes (data field 20002), which were assessed during the baseline interview, hospital inpatient records (HES; data field 41234) and primary care diagnosis records (data field 42040). For RS, code 1561 was used from the self-reported data. From the hospital inpatient data, we included individuals as a case for the phenotype if they had I73.0 ICD-10 or 4430 ICD-9 diagnosis code. In the primary care data, diagnoses are coded using the NHS-specific Read v2 or CTV3 codes instead of the ICD-coding. We used the following Read codes to define the respective phenotype:

- Read v2: “G730.”, “G7301”, “G7300” or “G730z”
- Read CTV3 (v3): “G730.”, “XE0VQ”, “G7300”, “G730z”, “G7301”, “X7051” or “XE0XA”

With this definition for RS, we ended up with 5,162 cases and 440,833 controls of European ancestry. Most of the cases for RS came from the primary care data (N = 2,953 (57.2%)) and hospital inpatient data (N = 1,664 (32.2%)).

Overall, of our RS cases 5,770 (50.8%) come from primary care data, 5,043 (44.4%) from hospital record data and 545 (4.8%) from self-reported data. Individuals who did not have the codes mentioned above were used as controls.

#### GWAS

For the FinnGen cohort, GWAS was conducted using the REGENIE (v.2.2.4) pipeline for R10 data^68^ (https://github.com/FINNGEN/regenie-pipelines). Analysis was adjusted for age at death or end of follow up (12.31.2021), sex, genotyping batches and the first 10 genetic principal components. Firth approximation was applied for variants with association P-value < 0.01.

The UKB GWA analyses were performed using REGENIE v3.1.1^68^. The whole-genome regression model (step 1) was created using 524,307 high-quality genotyped SNPs (bi-allelic; MAF ≥ 1%; HWE P > 1×10^-06^; present in all genotype batches, total missingness < 1.5% and not in a region of long-range LD^69^ with the leave-one-chromosome-out (--loocv) option enabled. We corrected for the following covariates:

- age at follow-up end (2019.08.18) or death (if earlier than follow-up end), calculated as the difference in years between the 15th day of month and year of birth (data fields 52 and 34, respectively) and the follow-up end or death date.
- sex (data field 31)
- genotyping array (categorical), derived from genotyping batch (data field 20000), as “UKB BiLEVE” (batches -11 to -1), “UKB Axiom release 1” (1 to 22) and “UKB Axiom release 2” (23 to 95).
- genetic principal components 1 to 10 (data field 22009)
- centre of baseline visit (categorical; data field 54)

The GWA (step 2) was performed using v3 imputed genotypes^58^ for chromosomes 1-22 and X with the approximate Firth correction applied for variants with association P-value < 0.05 (default setting), using the flags --firth, --approx and --firth-se. After analysis with REGENIE, we excluded results for imputed variants with MAF < 0.1% and/or imputation INFO < 0.3.

Association analysis in the EstBB was carried out for all variants with an INFO score > 0.4 using the additive model as implemented in REGENIE v3.0.3 with standard binary trait settings^68^. Logistic regression was carried out with adjustment for current age, age², sex and 10 first genetic principal components as covariates, analyzing only variants with a minimum minor allele count of 2.

In the MGB, the association analysis was carried out using REGENIE v3.2.2^68^, with covariates of Age, Sex, Genotype-Chip and five first principal components of ancestry calculated on just the analysis set (EUR only) of samples, analyzing only variants with a minimum minor allele count of 10.

Manhattan-plots for all the cohorts’ GWASs and meta-analyses were plotted using R version 4.0.1 (packages: qqman and RColorBrewer).

#### Meta-analysis

Meta-analyses were conducted using METAL (https://genome.sph.umich.edu/wiki/METAL_Documentation) with standard settings, and tracking allele frequency with the AVERAGEFREQ option and analyzing heterogeneity between used summary statistics with the ANALYZE HETEROGENEITY option. Summary statistics from different cohorts were matched against rsIDs. Both sample size based meta-analysis and effect estimate based analyses were run (Figure 1, Table 1 & Suppl. Table 3). Locus zoom plots from the meta-analysis results were created using the LocusZoom web browser (https://github.com/statgen/locuszoom)^70^.

#### eQTL and co-localization analyses

We conducted the eQTL analysis by using the web browser of the GTEx project (https://gtexportal.org/home/)^71^. Co-localization analyses were performed using the coloc R package (v5.1.0.1)^72,73^ in R v4.2.2. We extracted all variants in a 200kb region centered on the lead variant and imported the same region from GTEx v8^71^ eQTL association statistics. We then tested the co-localization between RS and each of the 49 tissues and generated co-localization plots using the LocusCompareR R package (v1.0.0)^74^ using LD r^2^ from 1000 Genomes European-ancestry samples.

### Functional assays

#### RNA extraction and RT-PCR

RNA was isolated according to manufacturer’s instructions using RNeasy plus micro kit (Qiagen, #74034). The quality of the RNA was determined with Nanodrop ND-1000 (Thermo Fisher Scientific), and 500 ng of total RNA was used for cDNA synthesis using High-capacity RNA-to-cDNA kit (Life Technologies, #4388950) on a BIO-RAD C1000 thermal cycler. RT-qPCR was performed using Taqman probes for *ADRA2A* (Hs01099503), *ADRA2C* (Hs03044628) and *NOTCH3* (Hs01128537) according to the manufacturer’s instructions on a ViiA7 Real-Time PCR system (Applied Biosystems, Foster City, CA). GAPDH and UBC were used to normalize relative expression levels. The 2–ΔΔCt method was used to quantify relative gene expression levels. Technical triplicates of Ct values were averaged for each sample and normalized to the housekeeping gene. Expression levels of mRNA are presented as fold change (control group = 1).

#### SiRNA and lentiviral overexpression

For siRNA transfection, cells were 60% confluent when treated with siRNA or scramble control to a final concentration of 20 nM with RNAiMax (Invitrogen, Carlsbad, CA). The siRNAs for *ADRA2A* (Cat # L-005422-00-0005) and *ADRA2C* (Cat # L-005424-00-0005) were purchased from Dharmacon (ONTARGET plus SMART pool siRNA). Cells were treated with an equimolar combination of Silencer and Scramble and collected 72 hours after transfection.

For overexpression studies *ADRA2A* (Human Tagged ORF Clone in pLenti-C-Myc-DDK Lentiviral Gene Expression Vector, NM_000681) and *ADRA2C* (Human Tagged ORF Clone in pLenti-C-Myc-DDK Lentiviral Gene Expression Vector, NM_000683) plasmids were purchased from OriGene Technologies. To package viruses 8.5×10^5^ HEK293T cells plated in each well of a six-well plate. The following day, lentiviral gene expression vectors were co-transfected with second generation lentivirus packaging plasmids, pMD2.G and pCMV-dR8.91, into cells using Lipofectamine 3000 (Thermo Fisher, L3000015) according to the manufacturer’s instructions. ViralBoost Reagent (AllStem Cell Advancements, VB100) was added (1:500) with fresh media after 5 hours. Supernatant containing viral particles was collected 48 hours after transfection and filtered. PASMCs were transduced with high MOI and treated for 12 hours and collected 72 hours after transfection.

#### RNAscope

Frozen sections of human coronary arteries were processed according to the manufacturer’s instructions, and all reagents were obtained from ACD Bio (Newark, CA). Sections were incubated with commercially available probes against human *ADRA2A* (#602791). Colorimetric assays were performed per the manufacturer’s instructions.

#### Single-cell RNA sequencing (scRNAseq) from human tissue

scRNAseq data was obtained from the human vascular atlas available on https://cellxgene.cziscience.com/.

#### Smooth muscle cell contraction assay

Pulmonary arterial SMCs were transfected with either scrambled siRNA or siADRA2A, siADRA2C or combination of both. Following 48 hours of transfection, cells were trypsinized and collected to be used for a collagen-based cell contraction assay Cyto-Select 48-well Cell Contraction Assay Kit (Cell Biolabs, San Diego, CA). In the assay, a mixture the pulmonary SMCs (3×10^6^/ml) and cold Collagen Gel Working Solution was incubated in a 48-well dish at 37°C for 1 hour to induce optimal polymerization following the manufacturer’s instructions. Next, cell culture medium with added SMC contraction agent or without was added on top of each well already containing the polymerized cell and collagen mixture. The cells were then incubated at either 28 °C or at 37°C, 5% CO_2_.

After 48 hours, the Keyence slide scanning microscope BZ-X810 was used to image the wells and cell contraction was measured using ImageJ by drawing the outlines of the gel, calculating the gel area, and comparing it to the well area.

#### Generation and analysis of CRIPSR lines

Genome editing of the region around rs7090046 was performed by CRISPRi/dCas9-KRAB system as previously reported^74^. The guide RNAs targeting this SNP were designed using Benchling online tools. Synthesized oligos were then cloned into pBA904 vector backbone containing dCas9-KRAB and lentivurs was packaged as described above. For the CRISPR interference experiment, PASMCs cells were seeded into 6 well plate (8×10^5^ cells /well). The next day, cells were transduced with the virus for 12 hours with 8 μg/mL polybrene. The cells were cultured for an additional 5 days with medium change until RNA was extracted. GuideRNA sequences are listed in Supplementary Table 6.

#### Primary cell culture and sample processing

Primary human pulmonary artery smooth muscle cells (PASMC) obtained from Dr. Rabenovich were originally isolated from pulmonary arteries (< 1mm) harvested from unused donor lungs all obtained de-identified from the Pulmonary Hypertension Breakthrough Initiative (https://ipahresearch.org/phbi-research/). For the experiments, total RNA was isolated from confluent cells at passage 5 and 6.

#### Statistics and reproducibility for the functional studies

All statistical analyses were conducted using GraphPad Prism software version 9 (Dotmatics Inc). Difference between two groups were determined using an unpaired two-tailed Student’s t-test. Differences between multiple groups were evaluated by one-way analysis of variance (ANOVA) followed by Dunnett’s post-hoc test after the sample distribution was tested for normality. P-values < 0.05 were considered statistically significant. All error bars represent standard error of the mean. Number of stars for the P-values in the graphs: *** P < 0.001; ** P < 0.01; * P < 0.05. No statistical method was used to predetermine sample size, which was based on extensive prior experience with this model.

## Supporting information

Supplementary_file_RS_Tervi_et_al

Supplementary_table_7_RS_Tervi_et_al

Supplementary_table_8_RS_Tervi_et_al

## Data Availability

Individual-level data for can be accessed on successful application for cohorts used in this study. The FinnGen individual-level data may be accessed through applications to the Finnish Biobanks' FinnBB portal, Fingenious (www.finbb.fi). For the individual-level data of the UKB, applications can be made through the UKB portal at https://www.ukbiobank.ac.uk/enable-your-research/apply-for-access. For MGB, individual-level data are available from the Mass General Brigham Human Research Office/Institutional Review Board at Mass General Brigham (contact located at https://www.partners.org/Medical-Research/Support-Offices/Human-Research-Committee-IRB/Default.aspx) for researchers who meet the criteria for access to confidential data. Lastly, for the EstBB, preliminary inquiries to access individual-level data for scientific research can be sent to.
Summary-level data is available upon reasonable request.

## Data and code availability

Individual-level data for can be accessed on successful application for cohorts used in this study. The FinnGen individual level data may be accessed through applications to the Finnish Biobanks’ FinnBB portal, Fingenious (www.finbb.fi). For the individual level data of theUKB, applications can be made through the UKB portal at https://www.ukbiobank.ac.uk/enable-your-research/apply-for-access. For MGB, individual level data are available from the Mass General Brigham Human Research Office/Institutional Review Board at Mass General Brigham (contact located at https://www.partners.org/Medical-Research/Support-Offices/Human-Research-Committee-IRB/Default.aspx) for researchers who meet the criteria for access to confidential data. Lastly, for the EstBB, preliminary inquiries to access individual level data for scientific research can be sent to.

Summary level data will be available upon publication in Dryad open-access repository.

## Conflicts of Interest

The authors declare no competing interests.

## Acknowledgments

A.T. received support for this work from the Instrumentarium Science Foundation (230041) and Doctoral Programme Brain and Mind (University of Helsinki). A.T. and H.M.O. received support from the Academy of Finland (340539). Additionally, M.R. received support from Sigrid Juselius Foundation, Emil Aaltonen foundation, Biomedicum Helsinki Foundation, Orion Research Foundation and from Finnish Foundation for Cardiovascular Research.

We want to acknowledge the FinnGen study and the FinnGen team for their contribution. We would like to thank the UK Biobank and the Mass General Brigham Biobank participants and staff for making this work possible. We also want to acknowledge the participants of the Estonian Biobank for their contributions. The Estonian Genome Center analyses were partially carried out in the High Performance Computing Center, University of Tartu.

The FinnGen project is funded by two grants from Business Finland (HUS 4685/31/2016 and UH4386/31/2016) and the following industry partners: AbbVie Inc., AstraZeneca UK Ltd, BiogenMA Inc., Bristol Myers Squibb, Genentech Inc., Merck Sharp Dohme Corp, Pfizer Inc., Glaxo-SmithKline Intellectual Property Development Ltd., Sanofi US Services Inc., Maze TherapeuticsInc., Janssen Biotech Inc, Novartis Pharma AG, and Boehringer Ingelheim. Following biobanks are acknowledged for delivering biobank samples to FinnGen: Auria Biobank (www.auria.fi/biopankki), THL Biobank (www.thl.fi/biobank), Helsinki Biobank (www.helsinginbiopankki.fi), Biobank Borealis of Northern Finland (https://www.ppshp.fi/Tutkimus-ja-opetus/Biopankki/Pages/Biobank-Borealis-briefly-in-English.aspx), Finnish Clinical Biobank Tampere (www.tays.fi/en-US/Research/and/development/Finnish/Clinical/Biobank/Tampere), Biobank of Eastern Finland (www.ita-suomenbiopankki.fi/en), Central Finland Biobank (www.ksshp.fi/fi-FI/Potilaalle/Biopankki), Finnish Red Cross Blood Service Biobank (www.veripalvelu.fi/verenluovutus/biopankkitoiminta) and Terveystalo Biobank (www.terveystalo.com/fi/Yritystietoa/Terveystalo-Biopankki/Biopankki/). All Finnish Biobanks are members of BBMRI.fi infrastructure (www.bbmri.fi) and FINBB biobank cooperative (https://finbb.fi/) is the coordinator of the BBMRI-ERIC operations in Finland covering all Finnish biobanks.

The work of the Estonian Genome Center, University of Tartu was funded by the European Union through Horizon 2020 research and innovation program under grants no. 810645 and 894987, through the European Regional Development Fund projects GENTRANSMED (2014-2020.4.01.15-0012), MOBEC008, MOBERA21 and Estonian Research Council Grant PRG1291

## Notes

### Competing Interest Statement

The authors have declared no competing interest.

### Funding Statement

This work was funded by the Instrumentarium Science Foundation (230041), the Academy of Finland (340539), Doctoral Programme Brain and Mind (University of Helsinki).

### Author Declarations

Ethics committee/IRB of the Institute for Molecular Medicine Finland gave ethical approval for this work. Ethics committee/IRB of the FinnGen study gave ethical approval for this work. Ethics committee/IRB of Stanford University gave ethical approval for this work. Ethics committee/IRB of the Estonian Biobank gave ethical approval for this work. Ethics committee/IRB of the Mass-General Brigham Biobank gave ethical approval for this work. Ethics committee/IRB of the UK Biobank gave ethical approval for this work.

### Summary of Updates

We updated the numbers of cases and controls for Mass-General Brigham Biobank and specified the origin of the data used (primary care versus hospital data) in our genetic analyses. In addition, some wording in the manuscript was improved.

