## Supplementary_file_RS_Tervi_et_al for "The α_2A_-adrenergic receptor (*ADRA2A*) modulates susceptibility to Raynaud’s syndrome"

### Supplementary Figures

#### Supplementary figure 1. Manhattan plots from all used cohorts

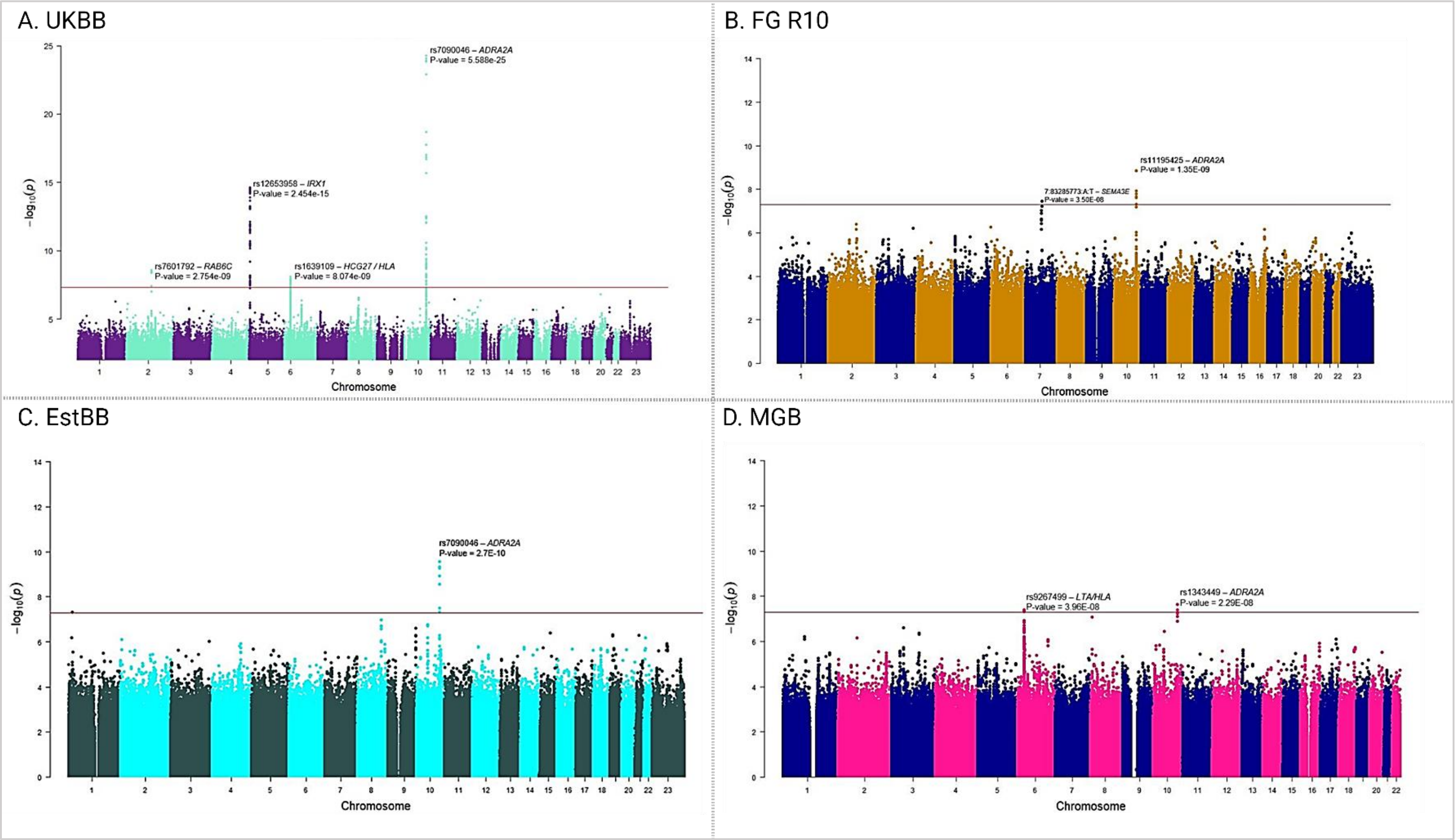

**Suppl.Fig.1.** Manhattan plots from GWA analyses A. UK Biobank, B. FinnGen data freeze 10, C. the Estonian Biobank, and D. the Mass-General Brigham Biobank.

### Supplementary figure 2. Manhattan plots UKB and FinnGen R10 – females and males

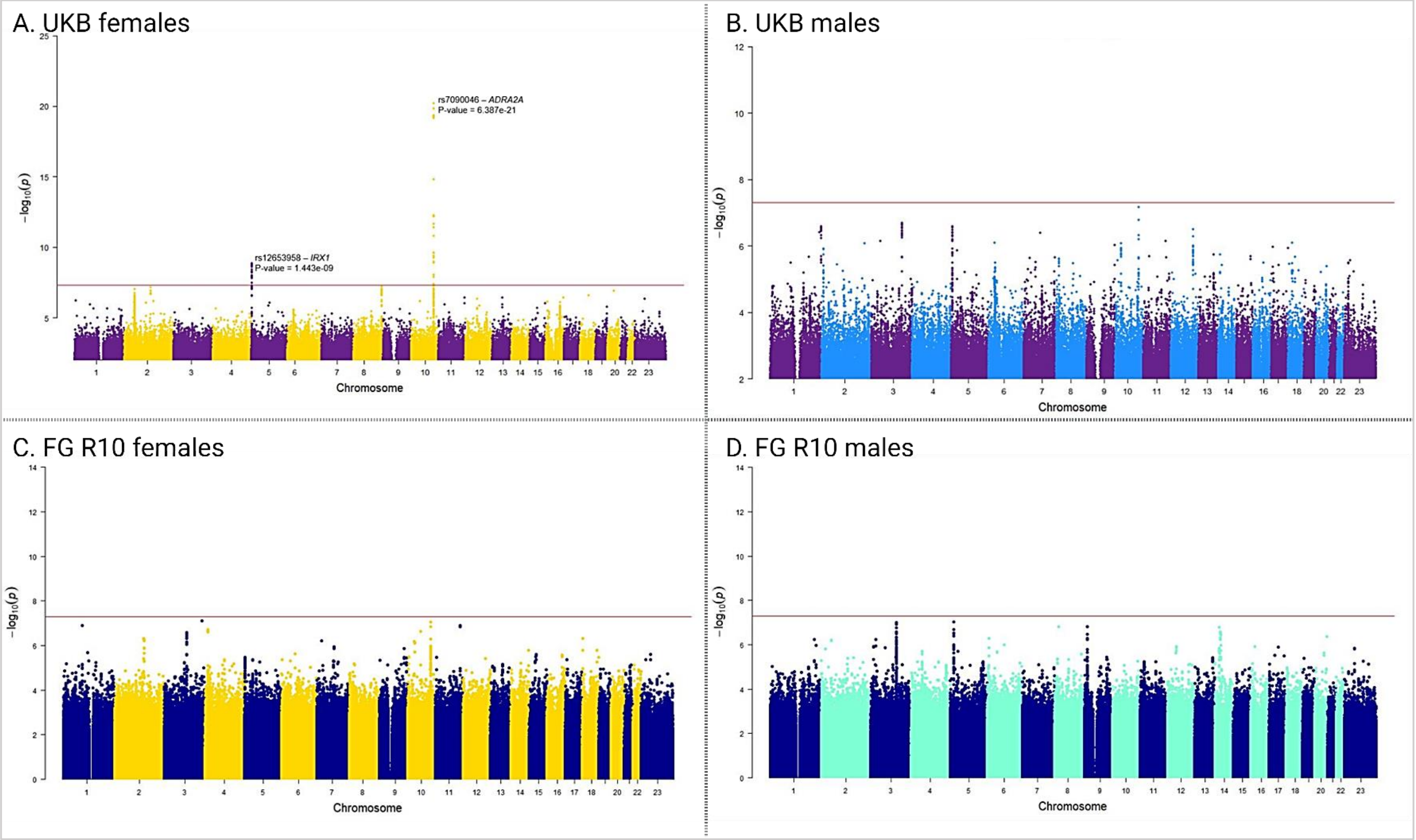

**Suppl.Fig.2.** Manhattan plots from sex-specific GWA analyses A. UK Biobank females, B. UK Biobank males, C. FinnGen data freeze 10 females, and D. FinnGen data freeze 10 males.

#### Supplementary figure 3. Co-localization plot for rs1343449

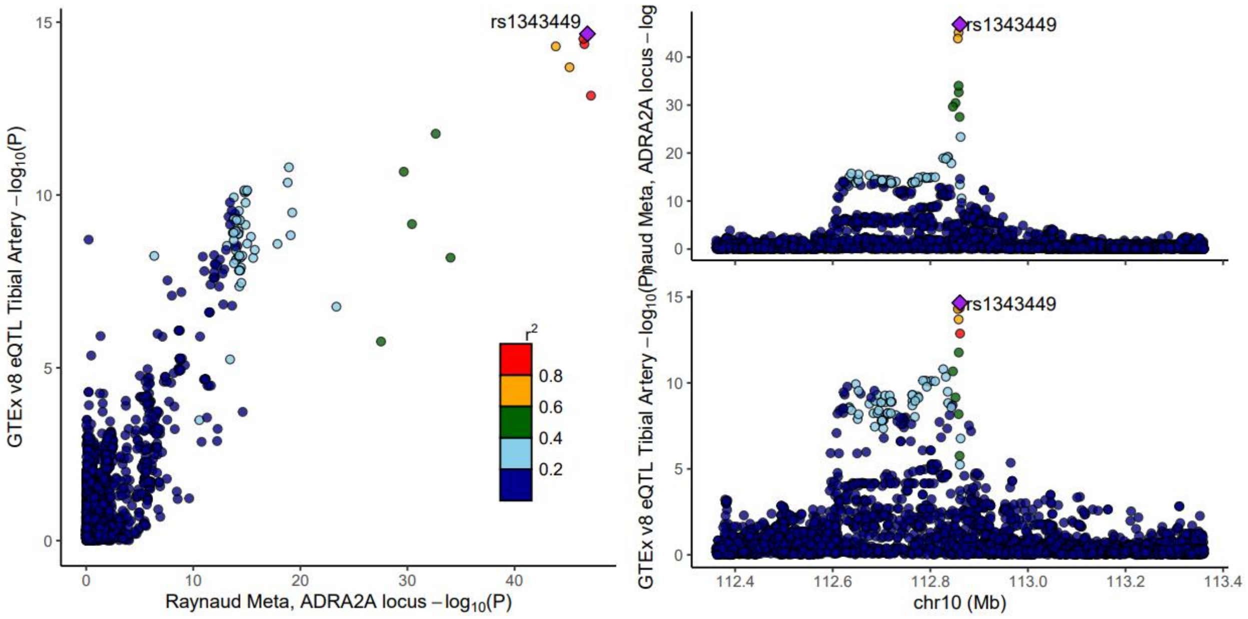

**Suppl.Fig.3.** Raynaud's syndrome association colocalizes with the lead variant for *ADRA2*A expression in GTEx in tibial arteries (rs1343449).

### Supplementary Tables

#### Supplementary table 1. Cohort demographics

| Cohort | RS cases | RS controls | Mean age of onset [SD] | Females in cases (%) |
|---|---|---|---|---|
| UKBB | 5,162 | 440,833 | 54.73 [14.13] | 3,691 (71.5%) |
| FinnGen R10 | 2,084 | 410,148 | 51.14 [16.20] | 1,446 (69.4%) |
| Estonia Biobank | 2,205 | 203,345 | 43.03 [15.37] | 1,672 (75.8%) |
| Mass General Brigham | 1,907 | 52,545 | *NA | 1,507 (79.0%) |
| Combined | 11,358 | 1,106,871 | 49.63 [15.23] | 8,510 (73.2%) |

*Information is not available for this cohort

Supplementary table 2. Cohort specific lead variants

**FinnGen R10 top 5 variants**

| SNP | CHR | POS (b38) | Ref. | Alt. | Combined Alt.AF | P-value | Beta | SE |
|---|---|---|---|---|---|---|---|---|
| rs11195425 | 10 | 111100270 | A | T | 0.129 | $1.35 \times 10^{-09}$ | 0.266 | 0.044 |
| rs7090046 | 10 | 111101172 | G | A | 0.247 | $1.19 \times 10^{-08}$ | 0.201 | 0.035 |
| rs1343451 | 10 | 111100633 | A | G | 0.247 | $1.58 \times 10^{-08}$ | 0.199 | 0.035 |
| rs1343449 | 10 | 111100768 | A | G | 0.247 | $2.12 \times 10^{-08}$ | 0.197 | 0.035 |
| rs7084501 | 10 | 111099941 | A | G | 0.248 | $2.50 \times 10^{-08}$ | 0.196 | 0.035 |

**UKB top 5 variants**

| SNP | CHR | POS (b37) | Ref. | Alt. | Combined Alt.AF | P-value | Beta | SE |
|---|---|---|---|---|---|---|---|---|
| rs7090046 | 10 | 112860930 | G | A | 0.310 | $5.59 \times 10^{-25}$ | 0.217 | 0.021 |
| rs7084501 | 10 | 112859699 | A | G | 0.314 | $8.65 \times 10^{-25}$ | 0.216 | 0.021 |
| rs1343449 | 10 | 112860526 | A | G | 0.316 | $1.27 \times 10^{-24}$ | 0.214 | 0.021 |
| rs7075626 | 10 | 112857971 | G | C | 0.333 | $1.39 \times 10^{-24}$ | 0.214 | 0.021 |
| rs1343451 | 10 | 112860391 | A | G | 0.314 | $1.50 \times 10^{-24}$ | 0.214 | 0.021 |

**The EstBB top 5 variants**

| SNP | CHR | POS (b37) | Ref. | Alt. | Combined Alt.AF | P-value | Beta | SE |
|---|---|---|---|---|---|---|---|---|
| rs7090046 | 10 | 112860930 | G | A | 0.234 | $2.68 \times 10^{-10}$ | 0.227 | 0.036 |
| rs1343449 | 10 | 112860526 | A | G | 0.235 | $4.72 \times 10^{-10}$ | 0.223 | 0.036 |
| rs7084501 | 10 | 112860391 | A | G | 0.239 | $4.95 \times 10^{-10}$ | 0.222 | 0.036 |
| rs1343451 | 10 | 112859699 | A | G | 0.235 | $5.25 \times 10^{-10}$ | 0.222 | 0.036 |
| rs7075626 | 10 | 112857971 | G | C | 0.269 | $1.17 \times 10^{-09}$ | 0.209 | 0.034 |

**The Mass-General Brigham Biobank top 5 variants**

| SNP | CHR | POS (b38) | Ref. | Alt. | Combined Alt.AF | P-value | Beta | SE |
|---|---|---|---|---|---|---|---|---|
| rs1343449 | 10 | 111100768 | A | G | 0.329 | $2.29 \times 10^{-08}$ | 0.186 | 0.033 |
| rs9267499 | 6 | 31569519 | G | C | 0.105 | $3.96 \times 10^{-08}$ | -0.297 | 0.056 |
| rs1343451 | 10 | 111100633 | A | G | 0.334 | $4.06 \times 10^{-08}$ | 0.181 | 0.033 |
| rs4947324 | 6 | 31560353 | C | T | 0.105 | $4.82 \times 10^{-08}$ | -0.295 | 0.056 |
| rs7084501 | 10 | 111099941 | A | G | 0.331 | $4.97 \times 10^{-08}$ | 0.181 | 0.033 |

Supplementary table 3. Sample size meta-analysis UKB, FinnGen R10, the Estonian Biobank and the Mass-General Brigham Biobank lead variants

| SNP | CHR | POS (b38) | Ref. | Alt. | P-value | Heterogeneity P-value | Locus name |
|---|---|---|---|---|---|---|---|
| rs7090046 | 10 | 111101172 | G | A | $4.08 \times 10^{-44}$ | 0.001 | *ADRA2A* |
| rs56324263 | 5 | 4045532 | C | T | $8.21 \times 10^{-23}$ | 0.028 | *IRX1* |
| rs3130968 | 6 | 31097294 | C | T | $8.11 \times 10^{-12}$ | 0.244 | *C6orf15/HLA* |
| rs743507 | 7 | 151010400 | C | T | $1.79 \times 10^{-09}$ | 0.172 | *NOS3* |
| rs6988032 | 8 | 50580957 | G | T | $5.28 \times 10^{-08}$ | 0.201 | *SNTG1* |

Supplementary table 4. Comparison of the lead variant in different cohorts

| SNP | FinnGen R10 | UK Biobank | The Estonian Biobank | The Mass General Brigham Biobank | Meta-analysis |
|---|---|---|---|---|---|
| rs7090046 | P = $1.19 \times 10^{-08}$<br>Beta = 0.201<br>SE = 0.035 | P = $5.59 \times 10^{-25}$<br>Beta = 0.217<br>SE = 0.021 | P = $2.40 \times 10^{-11}$<br>Beta = 0.260<br>SE = 0.038 | P = $5.84 \times 10^{-08}$<br>Beta = 0.181<br>SE = 0.033 | P = $7.32 \times 10^{-48}$<br>Beta = 0.213<br>SE = 0.015 |

Supplementary table 5. ENCODE stratified LD score regression

| Name | Coefficient | Coefficient_std_error | Coefficient_P_value |
|---|---|---|---|
| **Stomach_Smooth_Muscle__H3K4me1** | $8.40 \times 10^{-09}$ | $2.77 \times 10^{-09}$ | $1.20 \times 10^{-03}$ |
| **Stomach_Smooth_Muscle__H3K27ac** | $6.04 \times 10^{-09}$ | $2.11 \times 10^{-09}$ | $2.12 \times 10^{-03}$ |
| **Rectal_Smooth_Muscle__H3K4me1** | $6.99 \times 10^{-09}$ | $2.51 \times 10^{-09}$ | $2.70 \times 10^{-03}$ |
| **Duodenum_Smooth_Muscle__H3K4me1** | $8.65 \times 10^{-09}$ | $3.16 \times 10^{-09}$ | $3.08 \times 10^{-03}$ |
| **Fetal_Stomach__H3K4me1** | $7.27 \times 10^{-09}$ | $2.66 \times 10^{-09}$ | $3.09 \times 10^{-03}$ |
| **Esoph-Mucosa_ENTEX__H3K4me1** | $4.76 \times 10^{-09}$ | $1.89 \times 10^{-09}$ | $5.94 \times 10^{-03}$ |
| **Artery-Coronary_ENTEX__H3K27ac** | $5.51 \times 10^{-09}$ | $2.31 \times 10^{-09}$ | $8.46 \times 10^{-03}$ |
| **Stomach_Smooth_Muscle__H3K4me3** | $1.21 \times 10^{-08}$ | $5.15 \times 10^{-09}$ | $9.49 \times 10^{-03}$ |
| **Vagina_ENTEX__H3K4me3** | $1.58 \times 10^{-08}$ | $6.80 \times 10^{-09}$ | 0.010 |
| **Spleen__H3K4me3** | $1.29 \times 10^{-08}$ | $5.58 \times 10^{-09}$ | 0.011 |
| **NHDF-Ad_Adult_Dermal_Fibroblast_Primary_Cells__DNase** | $9.51 \times 10^{-09}$ | $4.16 \times 10^{-09}$ | 0.011 |
| **Fetal_Muscle_Trunk__H3K4me1** | $4.05 \times 10^{-09}$ | $1.84 \times 10^{-09}$ | 0.014 |
| **Vagina_ENTEX__H3K27ac** | $3.87 \times 10^{-09}$ | $1.76 \times 10^{-09}$ | 0.014 |
| **Fetal_Stomach__H3K27ac** | $6.66 \times 10^{-09}$ | $3.04 \times 10^{-09}$ | 0.014 |
| **Rectal_Smooth_Muscle__H3K27ac** | $4.24 \times 10^{-09}$ | $1.96 \times 10^{-09}$ | 0.015 |
| **Aorta__H3K4me3** | $2.03 \times 10^{-08}$ | $9.66 \times 10^{-09}$ | 0.018 |
| **Esoph-Mucosa_ENTEX__H3K36me3** | $6.09 \times 10^{-09}$ | $3.00 \times 10^{-09}$ | 0.021 |
| **Foreskin_Fibroblast_Primary_Cells_skin02__H3K4me1** | $4.10 \times 10^{-09}$ | $2.04 \times 10^{-09}$ | 0.022 |

Supplementary table 6. GuideRNA sequences targeting rs7090046 in CRISPRi analysis

| Position | Strand | Sequence | PAM | Specificity Score | Efficiency Score |
|---|---|---|---|---|---|
| **111101236** | 1 | CACCGggcgtagtgactgatgtgat | ggg | 76.2198801 | 68.94539763892847 |
| **111101150** | 1 | CACCGctacaattgaggaaacagaa | agg | 41.2282928 | 65.98685688262587 |
| **111101243** | 1 | CACCGtgactgatgtgatgggtcag | ggg | 58.2509063 | 65.63862659271983 |
